# Urban Vulnerability Assessment for Pandemic surveillance: The COVID-19 case in Bogotá, Colombia

**DOI:** 10.1101/2020.11.13.20231282

**Authors:** Jeisson Prieto, Rafael Malagón, Jonatan Gomez, Elizabeth León

## Abstract

A Pandemic devastates the life of global citizens and causes significant economic, social, and political disruption. Evidence suggests that Pandemic’s likelihood has increased over the past century because of increased global travel and integration, urbanization, and changes in land use. Further, evidence concerning the urban character of the Pandemic has underlined the role of cities in disease transmission. An early assessment of the severity of infection and transmissibility can help quantify the Pandemic potential and prioritize surveillance to control of urban areas in Pandemics. In this paper, an Urban Vulnerability Assessment (UVA) methodology is proposed. UVA investigates the possible vulnerable factors related to Pandemics to assess the vulnerability in urban areas. A vulnerability index is constructed by the aggregation of multiple vulnerability factors computed on each urban area (i.e., urban density, poverty index, informal labor, transmission routes). UVA provides insights into early vulnerability assessment using publicly available data. The applicability of UVA is shown by the identification of high-vulnerable areas where surveillance should be prioritized in the COVID-19 Pandemic in Bogotá, Colombia.

## 1. Introduction

Pandemics are large-scale outbreaks of infectious diseases that increase morbidity and mortality over a big geographic area and cause significant social, political, and economical disruption [1,2]. Previous Pandemics [1], have exposed gaps related to the timely detection of disease, tracing of contacts, availability of basic care, quarantine and isolation procedures, and health sector preparedness (i.e., global coordination and response mobilization) [3,4]. Suddenly, significant policy attention has focused on the need to identify and limit emerging outbreaks that might lead to Pandemics and to expand and sustain investment to build preparedness and health capacity [5]. Nonetheless, the timeliness of implementing these measures is paramount to control a highly contagious disease. Efficient prioritization of investigation of high-vulnerable areas would optimize the use of resources and potentially limit the size of the Pandemic [6–8].

Vulnerability Assessment describes the degree to which socioeconomic systems and physical assets in geographic areas are either susceptible or resilient to the impact of disaster (i.e., Pandemic). Once priority vulnerable areas are identified, it will be possible to understand why certain locations may need to be prioritized for preventative action and response efforts (i.e., planning and coordination, reducing the spread of disease, continuity of health care provision) [1,2,9,10]. In the urban context, the Urban Vulnerability Assessment (UVA) helps to determine what types of preparedness and response activities might help for an optimal Urban Strategic Planning (USP) to assist the decision-making processes undertaken by today’s urban planners [11]. Further, the recently published response plan for the current COVID-19 Pandemic, UN-Habitat has underlined the urban-centric character of the infectious disease [12]. It says, more than 1430 cities are affected by the Pandemic in 210 countries and well above 95% of the total cases are located in urban areas. Further, the World Health Organization (WHO) emphasized that the first transmission in the COVID-19 Pandemic did happen in the internationally connected megacities [13]. Even though there is urban universality of the disease, the cities of the global south are more susceptible given their population densities, low income, risky occupations, and lack of affordable health services [14].

Several models have been proposed to quantify vulnerable urban areas over the infectious disease domain, i.e., vector-borne diseases [15], Dengue [9], malaria [16,17], and Ebola [10]. More recently, in [18] a COVID-19 vulnerability index for urban areas in India is proposed, the vulnerability index aggregate weighted scores of a set of variables related to COVID-19 precaution of social distance and lockdown in four metro cities in India. However, an apriori knowledge or relative preferences between criteria based judgments for the gathering of preferences for indicators (vulnerable factors) is needed in those models. Different methodologies allow to transform the experts’ knowledge into a mathematical language (i.e., Analytic Hierarchy Process), but these methodologies have some limitations such as a-priori knowledge, expert bias, or hierarchical criteria [19,20].

In this paper, a conceptual framework for Urban Vulnerability Assessment (UVA) for Pandemics is proposed. UVA conducted a comprehensive review of relevant literature to identify vulnerable factors influencing Pandemics. These factors are used to generate an index that allows us to identify and rank potentially vulnerable urban areas. The rank is built using Borda’s count aggregation method, which does not need experts knowledge nor additional parameters for the construction of the ranking. Then, the vulnerability rank is associated with a vulnerability index, i.e., a higher rank indicates higher vulnerability. UVA is tested in the current COVID-19 Pandemic in Bogotá, the crowdest city of Colombia. Using widely available data of Bogotá (i.e., from the National Department of Statistics (DANE), the District Planning Secretary (SDP), and the District Mobility Secretary (SDM)), UVA creates a spatially explicit COVID-19 vulnerability index of Bogotá. To our knowledge, our study is the first to develop a composite measure of community-level vulnerability concerning the COVID-19 situation in Bogotá. The main value of our study is the Urban sector ranking provided to policymakers to prioritize resource allocation and devise effective mitigation and reconstruction strategies for affected populations in Bogotá.

This paper is divided into four sections. Section 2 develops the methodology of Urban Vulnerability Assessment (UVA) for Pandemic surveillance. Section 3 describes the applicability of UVA for the current COVID-19 Pandemic in Bogotá, Colombia. Finally, section 4 discusses some of the conclusions and potential future developments.

## 2. Vulnerability assessment

The conceptual framework of Urban Vulnerability Assessment (UVA) for Pandemics is illustrated in Figure 1. UVA involves four main stages. The first stage is the identification of vulnerable factors influencing Pandemics, see Figure 1(a). The second stage is to transform the raw input data from each vulnerable factor into a probability distribution, see Figure 1(b). The third state groups geographic areas with similar characteristics into classes to assign a vulnerability level, see Figure 1(c). After that, an aggregation method is applied to create a unique rank for each class, see Figure 1(d), where a higher rank is assigned to a higher vulnerability level, see Figure 1(e).

**Figure 1.**
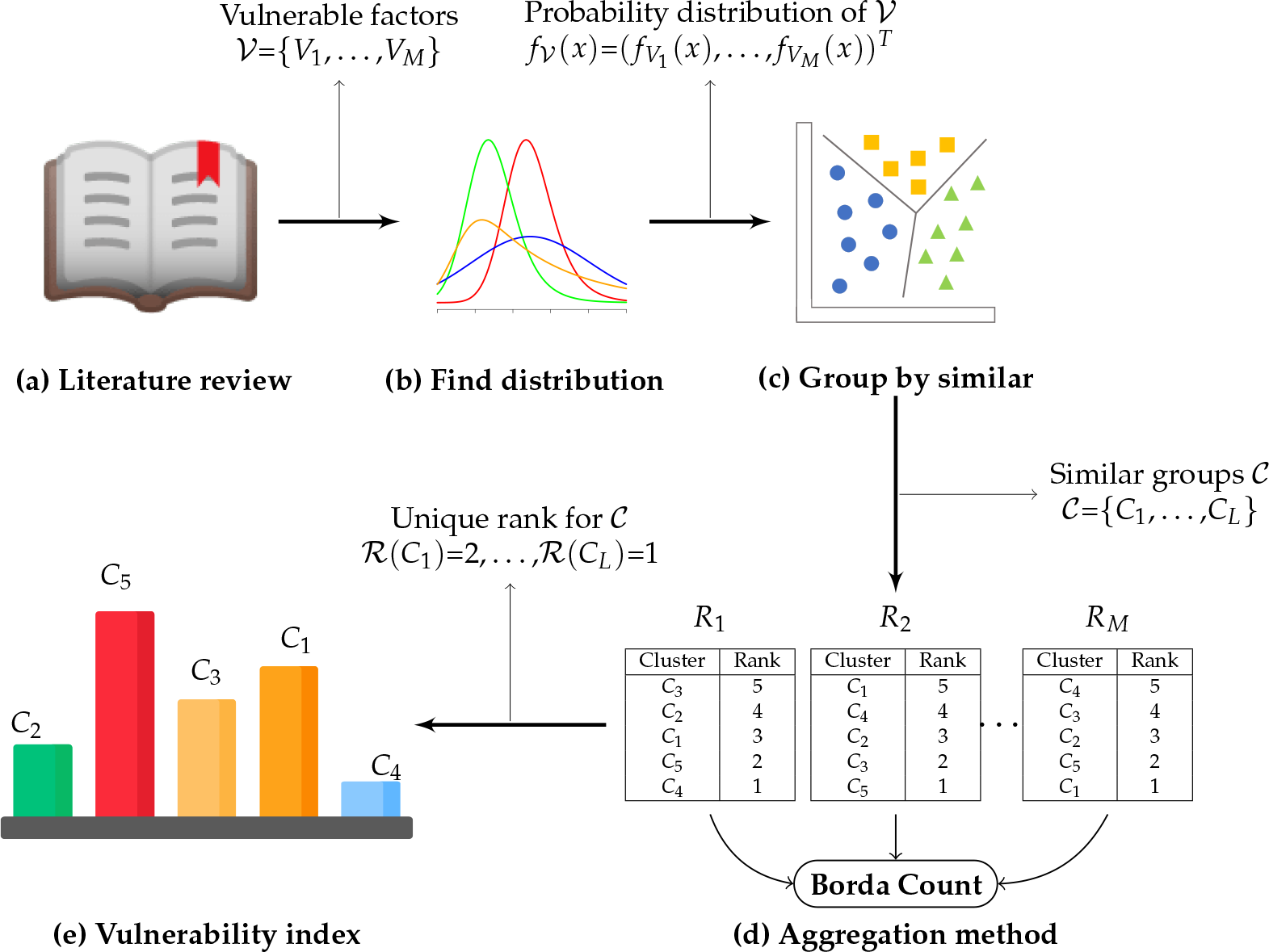
Schematic diagram of the urban vulnerability assessment in Pandemic disaster ^1^.

### 2.1. Literature review

We conducted a literature search to identify a set of peer-reviewed studies that possibly examined types of vulnerability factors related to Pandemics, see Figure 1(a). The studies consider both factors related to past Pandemics (i.e, 1881 Fifth cholera, 1918 Spanish flu influenza, 1957 Asian flu influenza, 2003 SARS, 2009 h1n1, 2013 West Africa Ebola) and factors found in the current COVID-19 Pandemic. The search retrieved studies for which the study’s title, abstract, or keywords indicated the study examined a type of vulnerability in Pandemics. Then, a manual assessment is made for every study against eligibility criteria:

- The study provided a quantitative or conceptual analysis of a type(s) of vulnerability factors related to infectious diseases (or Pandemics).
- The actual analysis or argument of the study earnestly included vulnerability.
- The study focuses on urban areas.
- To be eligible the study focuses more on the vulnerability analysis at geographic area than on the individual vulnerability of infectious diseases.

Then, the vulnerability factor the study focused on, the geographic focus of the study, and the methods used to assess the vulnerability is recorded. This involved examining the title, abstract, or keywords, or full-text version if required. We listed the country or region(s) where the study focused. For theoretical studies, studies that presented examples without a geographic focus, or studies with an unclear geographic focus, the geographic location is listed as Not Applicable (NA). Table 1 summarizes the 11 studies that were considered for the analysis of vulnerability factors related to Pandemics.

**Table 1.**
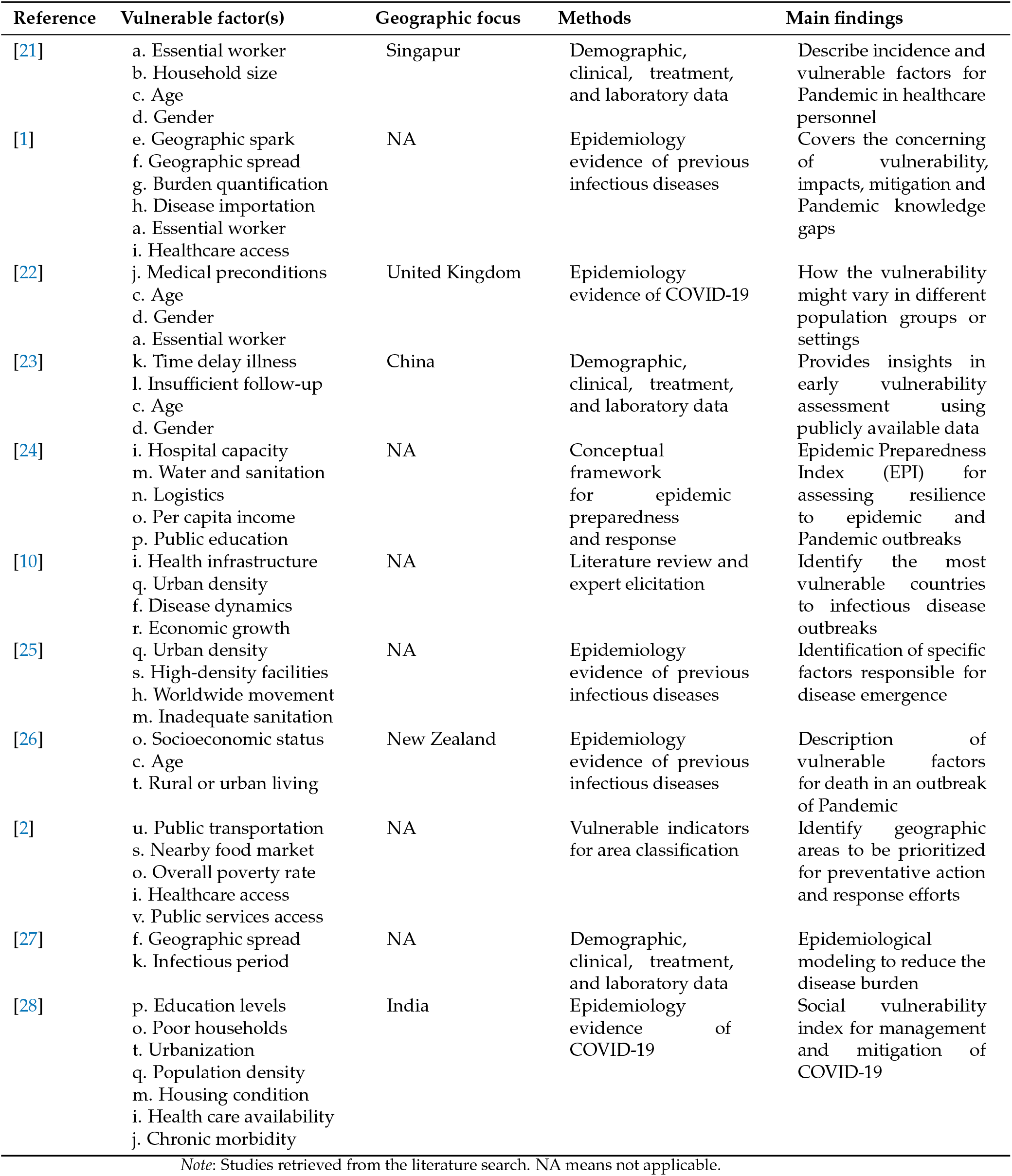
Summary of studies considered to vulnerability factors related with Pandemics.

### 2.2. Statistical data analysis

Let *𝒮* a geographical space under investigation (i.e. state, country, or city) defined in terms of a finite set of *N* smaller spatial units (i.e. countries, census tracts, or zip codes); that is *𝒮* = *{*1, 2, …, *N}*. Let *𝒱* a set of *M* vulnerable factors, and *V*_*k*_ the values of the *N* spatial units in the *k*-th vulnerable factor *V*_*k*_ = *{𝓋* _*k*,1_, …, *𝓋* _*k,N*_*}*. The raw data for each factor are normed across all spatial units over the range 0 (best) to 1 (worst), see Figure 1(b). Different normalization methods exists in the literature [29]. However, depending on the data properties, some normalization operations are inappropriate (i.e., anomalies, orthogonality, linear dependency). One solution is to build an estimation of the Probability Density Function (PDF) of the data, and then transform it via its Cumulative Density Function (CDF), so intervals with higher likelihood of containing data are assigned to higher portion of the normalized interval [0,1]. This is call probability integral transform [30]. We estimate the PDF 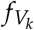 at specific spatial unit *x* using the Kernel Density Estimation (KDE) method.

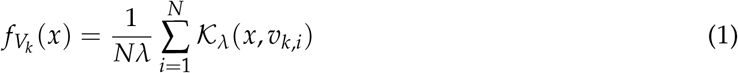

where *K* is the kernel (a non-negative function) and *λ* is the smoothing parameter called the *bandwith*.

Then, to normalize the raw data at spatial unit *x* over the range 0 (best) to 1 (worst) in the *k*-th vulnerable factor, the probability integral transform is applied.

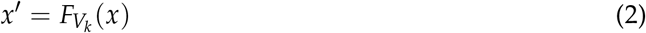

where 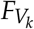 is the CDF of the *k*-th vulnerable factor.

### 2.3. Cluster Analysis

As a proposal to identify spatial units with possible high levels of vulnerability, a cluster analysis is made to group spatial areas with similar characteristics. Cluster analysis is a multivariate analysis technique that aims to organize information about variables (vulnerable factors) so that relatively homogeneous clusters can be formed i.e., synthesize the spatial units into *k* partitions. Therefore, each cluster consists of spatial areas with similar behavior from their PDF for the *M* vulnerable factors, see Figure 1(c).

UVA allows the decision-maker to select the number of *k* partitions in which the spatial units will be grouped. Each sub-set of solution *C* = *{C*_1_, …, *C*_*L*_*}* obtained by a cluster algorithm (i.e., k-means) contains a number *N*_*j*_ of spatial units of similar characteristics. In this way, the decision tool makes it possible to get an affordable number of *k* relevant possible vulnerable assessments (i.e., *k* = 3 vulnerability of low, medium, and high; *k* = 10 vulnerability from 1 to 10).

### 2.4. Create vulnerability index

In order to assign a vulnerability level (rank) to each cluster, a Borda’s count aggregation method is proposed [31]. The Borda’s method takes as input a set of ranks *R* = *{R*_1_, …, *R*_*M*_*}* (where *R*_*k*_ is an order of the Clusters *C* = *{C*_1_, …, *C*_*L*_*}* in the *k*-th vulnerability factor), and produce single rank by mixing the orders of all the input ranks. The number of points (weight) assigned for each ranking varies depending on which of several variants of the Borda count is used. For this, let 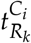 the position of the Cluster *C*_*i*_ in the rank *R*_*k*_, and 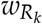 the weight assigned for the rank *R*_*k*_. A new aggregated value of ranking or the *i*-th Cluster is defined as:

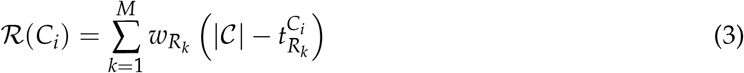

The rank built using Borda’s count aggregation method does not need experts knowledge nor additional parameters for the construction of the ranking (i.e., the weights could be equal for each rank). However, if the rank has to be weighted in some way, the method allows assigning this weight for each rank. For the vulnerability level assignation problem, a set of ordered lists or ranks is calculated using the *centroid* of each cluster *C*_*i*_. Therefore, Borda’s count aggregation method is used, that is, vulnerability factor ranks *R*_*k*_ were made sorted the values of each centroid for the *M* vulnerability factors. Next, these *M* ranks were combined using Borda’s method to construct the aggregated vulnerability rank, see Figure 1(d).

Finally, the vulnerability rank is associated with a vulnerability index, i.e., higher rank indicates higher vulnerability, see Figure 1(e).

## 3. Vulnerability index for the COVID-19 in Bogotá, Colombia

### 3.1. Study area and data sources

UVA is tested by the creation of a vulnerability index for the current COVID-19 Pandemic in Bogotá city, the largest and crowded city in Colombia. Bogotá is a metropolitan city with 7.412.566 inhabitants living in an area of 1775km (995km urban and 718km rural), at an altitude 2640m, with an annual temperature ranging from 6 to 20°C, and annual precipitation of over 840mm. Bogotá has composed of 621 Urban Sectors^2^. Each Urban sector belongs to one of the 112 Zonal Planning Units (UPZ), see Figure 2.

**Figure 2.**
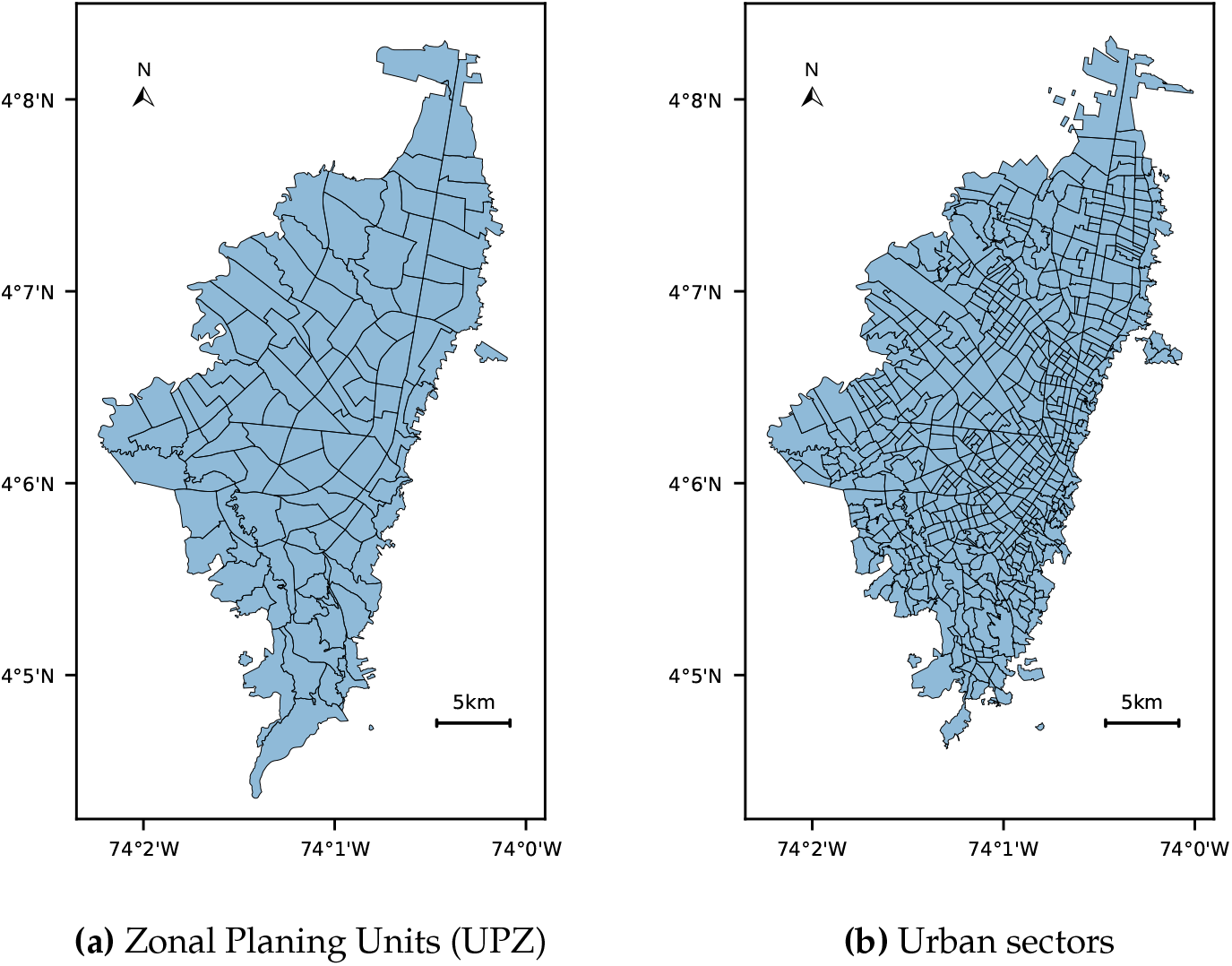
Spatial distribution of Bogotá, Colombia using Zonal Planing Units (UPZ) (left) and Urban sectors (right) ^3^.

Information was obtained from the National Department of Statistics (DANE), from District Planning Secretary of Bogotá (SDP), and District Mobility Secretary of Bogotá (SDM). Data comprised public information about demographic, transportation, socio-economic, and health conditions reported from 2011 to 2020. A summary of the dataset is presented as follows:

- **MON_2017 [32,33]:** Dataset provided by SDP and it contains set monographs to provide a physical, demographic and socioeconomic vision of Bogotá and its districts.
- **SDM_2017 [34]:** Dataset provided by SDM and it presents detailed official information of the characterization of mobility in Bogotá.
- **CNPV_2018 [35]**: Dataset provided by DANE and it is the national census made in 2018 and provides statistics about socio-demographic information Colombia.
- **DANE_2018 [36]:** Dataset provided by DANE and it contains the results of the Multidimensional Poverty Index that analyze educational conditions, health, work, access home public services, and housing conditions.
- **DANE_2020 [37]:** Dataset provided by DANE and it presents a vulnerability index based on demographic and health conditions relevant for COVID-19 Pandemic.

Since the datasets’ information are in different spatial units (i.e., Urban sectors, UPZ), the spatial unit using in this study is the Urban sector (more atomic), and the information at UPZ level is then transformed into Urban sectors by spatial transformation (i.e., a UPZ contains one (or more) Urban sectors, then the UPZ values are assigned to the Urban sector).

### 3.2. Vulnerability domains

Given the public data available for Bogotá, and the vulnerable factors find in the literature review (see Section 2.1) a set of domains is proposed to analyze the vulnerability in Bogotá. Throughout the course of the study, we found that the most-relevant concepts and associated measures fell into three common domains: (i) *Where and how he/she lives*, (ii) *Where and how he/she works*, and (iii) *Where and how he/she gets around* ^4^. Factors and associated measures within these three domains provide vulnerable factors for the quantitative analysis. Table 2 shows the domains proposed and their corresponding vulnerable factors associated with them.

**Table 2.**
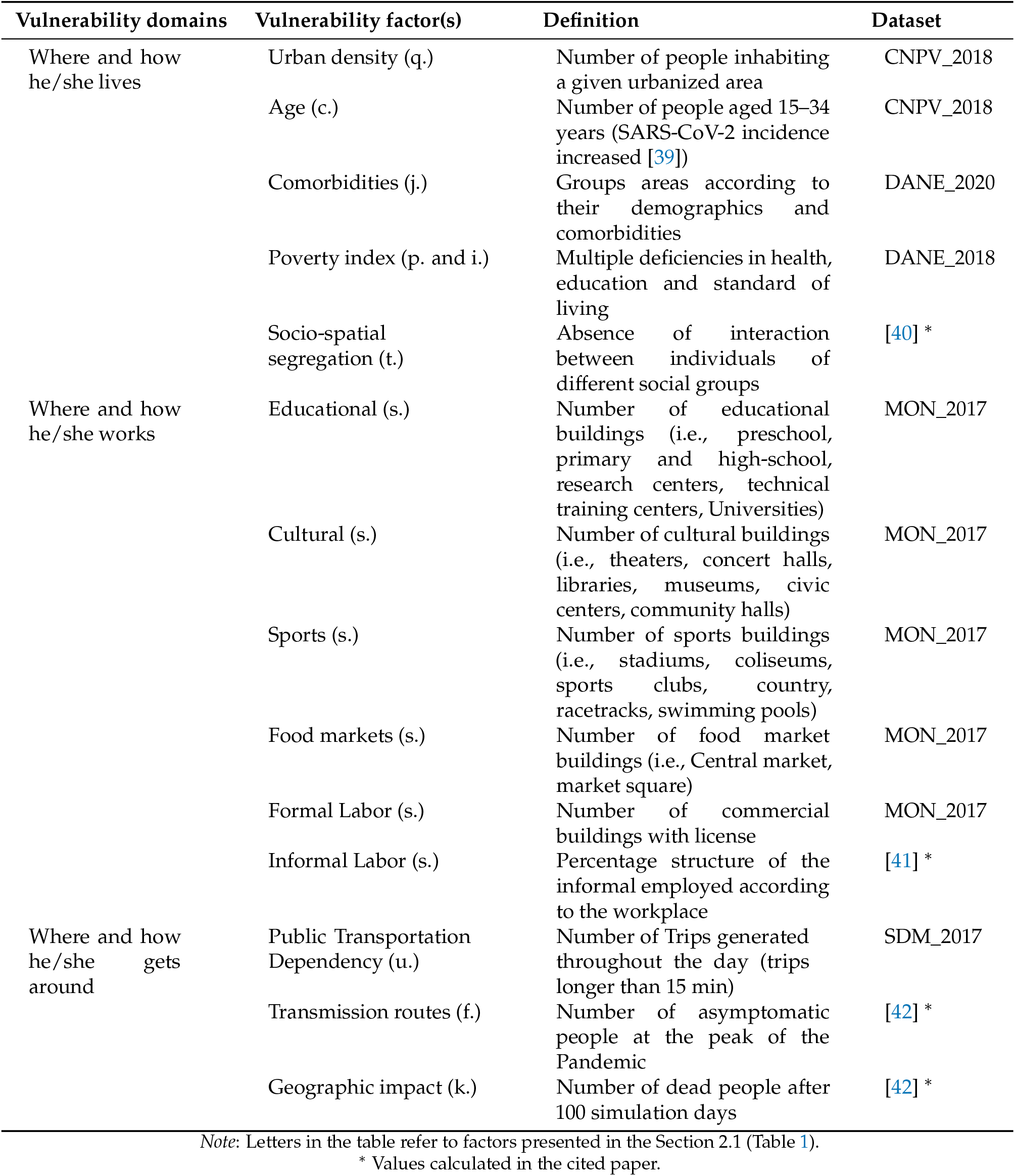
Vulnerability domains for the COVID-19 case in Bogotá, Colombia.

#### 3.2.1. Where and how he/she lives

Several demographic factors influence the degree of vulnerability of the Urban sector to Pandemics. The relevant literature emphasizes the role of such factors as *urban density, age*, and the urban living (i.e., *socio-spatial segregation*). The level of education or literacy, and the quality of the health care system (i.e., included in the *poverty index*) can also play a helpful role in mitigating the spread and effects of infectious diseases [10]. Further, most data on COVID-19 Pandemic suggest that people with underlying health conditions such as respiratory and cardiovascular disease, and cancer (i.e., *comorbidities*) are more vulnerable than people without them.

#### 3.2.2. Where and how he/she works

Urban sectors with high-density facilities (i.e, *educational buildings, cultural buildings, sport buildings, food markets*, all *formal labor*) are more vulnerable to the spread of contagious diseases due to space limitations within and between households, growth and mobility, and limited water, sanitation, and hygiene (WASH) infrastructure. Also, the overwhelming majority of workers in the informal economy (i.e., *informal labor*) have higher exposure to occupational health and safety vulnerability, no appropriate protection, forced to work for daily sustenance, and out-of-pocket costs [38].

#### 3.2.3. Where and how he/she gets around

Understand transmissibility, risk of geographic spread, *transmission routes*, and vulnerable factors for infection (i.e., *geographic impact*) provides the baseline for epidemiological modeling that can inform the planning of response and containment efforts to reduce the burden of disease [27]. As well, there have been claims that the use of public transport (i.e., *public transportation dependency*) has also led to the spread of infectious diseases [2]

### 3.3. Vulnerability analysis

To understand the distribution of the vulnerability factors over the Urban sectors, the raw data for each factor are normed across all Urban sectors over the 0 (less vulnerable) to 1 (most vulnerable). The normalization was made using the probability integral transformation, and to estimate the probability density (PDF) of each vulnerable factor, the Kernel Density Estimation (KDE) was used ^5^.

Figure 3 shows the normalization for the three domains. The vulnerability value for each factor is associated with the founded distribution. The results show the spatial correlation that exists for some vulnerable factors, especially for the *Where and how she/he works* domain. In contrast to the *Where and how she/he lives* domain, where the spatial correlation is not clear and the vulnerability is distributed across the geography area under study (Bogotá).

**Figure 3.**
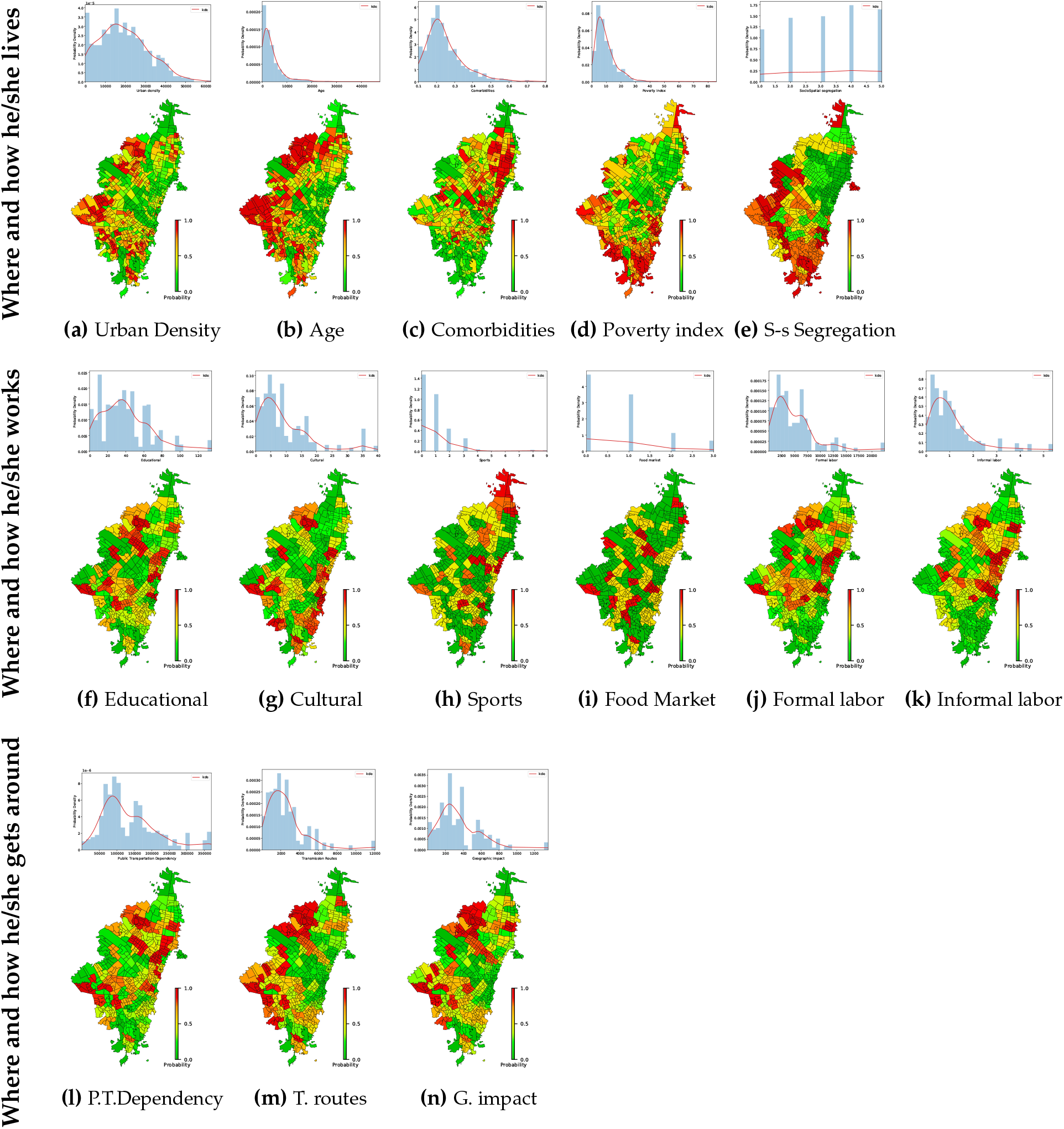
Probability density estimation for vulnerable factors using the KDE method.

### 3.4. Vulnerability index

To provide better response for vulnerability assessment, UVA generates three different vulnerability index to assess vulnerability in different ways (depending on the *k* partitions). Vulnerability index I have three different clusters (*k* = 3) to get a vulnerability index from low to high (i.e., low, medium, high). Vulnerability index II has five different clusters (*k* = 5) to get a vulnerability index from lowest to highest (i.e., lowest, low, medium, high, highest). And, Vulnerability index III has ten clusters (*k* = 10) to get a vulnerability index from 1 to 10. Figure 4 shows the different vulnerability index (i.e., Vulnerability index I with *k* = 3 (3 clusters), Vulnerability index II with *k* = 5 (5 clusters), Vulnerability index III with *k* = 10 (10 clusters)).

**Figure 4.**
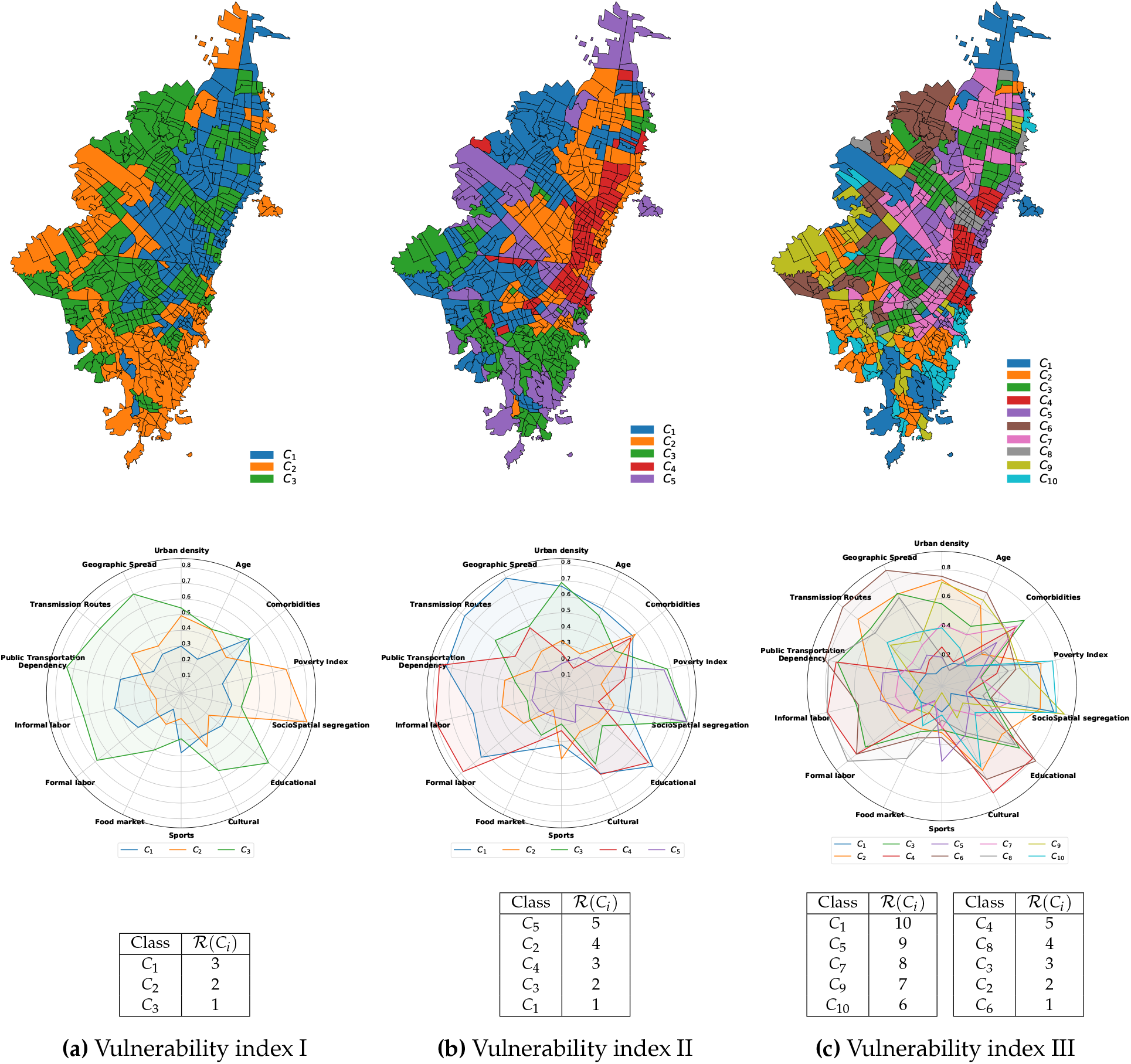
Three different proposed vulnerability indexes assess the urban Vulnerability. Vulnerability I with *k* = 3 (left panel), Vulnerability II with *k* = 5 (midst panel), Vulnerability III with *k* = 10 (right panel). For each Vulnerability index: clusters generated using the k-means method (top), its corresponding centroid values for each vulnerable factor (middle), and the unique rank generated using the Borda’s count method (bottom) ^6^.

After getting the clusters for each vulnerability index (showed in Figure 4 - top), the *centroids* of the clusters (showed in Figure 4 - midst) are getting and used to sort from higher to lower values the each vulnerability factor. These vulnerability factors sorted (by the *centroids*) are assumed as vulnerable ranks that would be used for the analysis. Then, to aggregate the 14 ranks (one for each vulnerable factor in Table 2) the Borda’s count aggregation method build a unique vulnerability ranking for each cluster (showed in Figure 4 - bottom).

A vulnerability index is assigned for each cluster based on the ranking (i.e., higher rank indicates higher vulnerability). Figure 5 shows the final vulnerability index for the three different vulnerability index constructed with UVA. For Vulnerability index I, the results show high vulnerable urban sectors in the south and west part of the city. On the other hand, the Vulnerability index II shows how some Urban sectors change from medium-vulnerability (in the Vulnerability index I) to low or high-vulnerability. The Vulnerability index III presents an interesting scenario where the spatial correlation between urban sectors is not remarkable getting a unbias vulnerability index for COVID-19. Although our intention was not to predict the risk of infection for an Urban sector, we observed similarities between vulnerability indexes proposed and the current concentration of COVID-19 cases confirmed in Bogotá, see Figure 6. The results show how vulnerable areas found with UVA match with urban areas with more COVID’19 cases. This indicates that the UVA framework proposed could be used to recommend actions for before, during, and after pandemic i.e., to planning and coordination efforts through leadership and coordination across sectors, to assess if the risk of a pandemic could increase in specific geographic areas.

**Figure 5.**
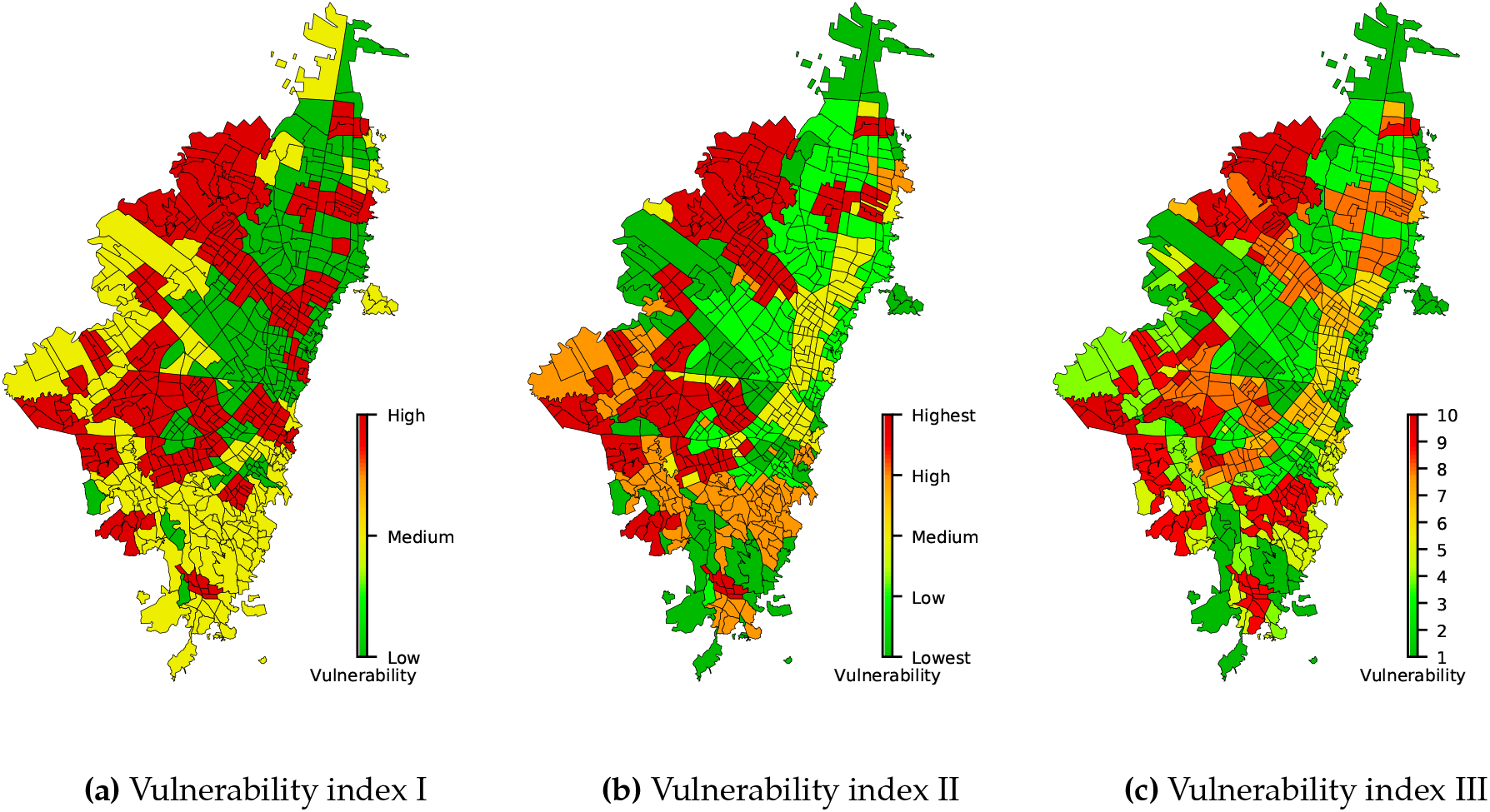
Vulnerability indices generated using UVA for the current COVID-19 Pandemic in Bogotá, Colombia. Vulnerability index I has 3 levels from low to high (left); Vulnerability index II has 5 levels from lowest to highest (middle); and Vulnerability index III has 10 levels from 1 to 10 (right).

**Figure 6.**
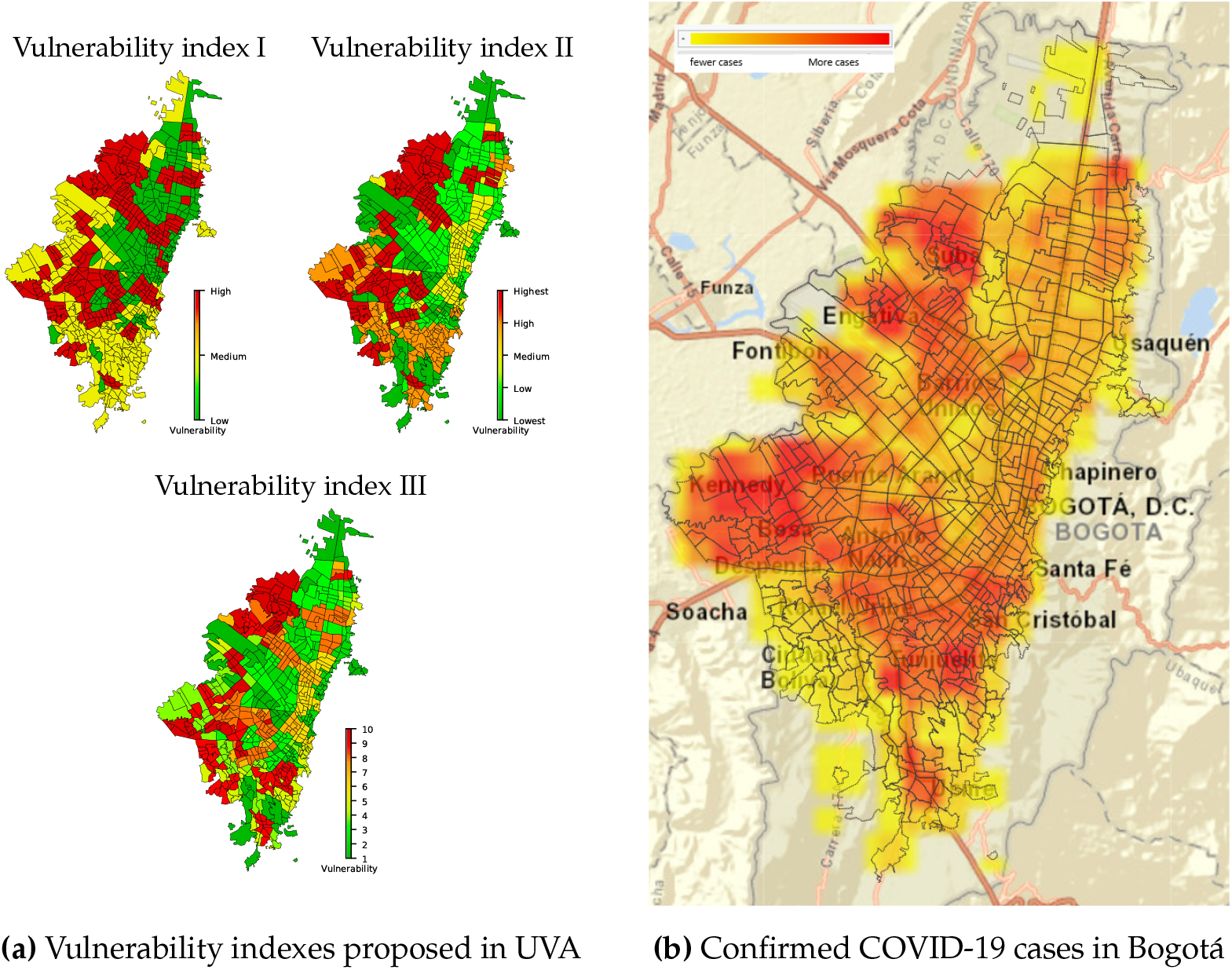
Comparison between the vulnerability indexes proposed in this study (left) and the real COVID-19 cases confirmed in Bogotá (right) ^7^. The colored boxes in the right map shows the concentrations of the cases in the Bogotá, where red color indicates more confirmed COVID-19 cases and fewer cases are in yellow.

## 4. Conclusions and Future Work

An Urban Vulnerability Assessment (UVA) for Pandemic surveillance is proposed. UVA highlights the vulnerability based on a set of 14 vulnerable factors found in the literature. The vulnerable factors are ranked for each spatial unit under study using some statistical methods like Kernel Density Estimation (KDE) and Clustering Analysis. To generate a unique vulnerability index, Borda’s count method is used. UVA is tested in the current COVID-19 Pandemic in Bogotá city, the largest and crowded city in Colombia. UVA creates not only one, but a set of vulnerability indices (i.e., low-high, lowest-highest, and 1-10) to Pandemic surveillance. Surveillance is of primary importance to monitor the burden of disease and will give both local authorities and the global community a chance for a quick response to public health threats.

Our work has demonstrated how high-vulnerable level contexts contribute to increasing the impact and spread of the disease at different geographic levels. This approach also enables the design of comprehensive plans, implemented at the city scale, for addressing urban vulnerability at national, regional, and provincial scales. Further, the results allow to build evidence for planning, modeling, and epidemiological studies to better inform the public, policymakers, and international organizations to where and how to improve surveillance, response efforts, and delivery of resources, which are crucial factors in containing the COVID-19 Pandemic. It must concern the spatial inequality problems in multiple deprivations and other depreciating characteristics. Thus enabling equity-based urban planning that vows to restrict the transmission of COVID-19 now or any similar Pandemic in the future.

## Data Availability

No medical data from external sources is used in this paper.
Data comprised public information about demographic, transportation, socio-economic, and health conditions reported from 2011 to 2020 of Bogotá, Colombia. Public information was obtained from the National Department of Statistics (DANE), from District Planning Secretary of Bogotá (SDP), and District Mobility Secretary of Bogotá (SDM).
DANE:
- https://www.dane.gov.co/files/comunicados/Nota_metodologica_indice_de_vulnerabilidad.pdf
- https://www.dane.gov.co/files/investigaciones/condiciones_vida/pobreza/2018/bt_pobreza_multidimensional_18.pdf
- http://microdatos.dane.gov.co/index.php/catalog/643
SDP:
- http://www.sdp.gov.co/sites/default/files/determinantes_de_la_distribucion_espacial_de_informalidad_laboral_en_bogota.pdf
- http://www.sdp.gov.co/gestion-estudios-estrategicos/informacion-cartografia-y-estadistica/repositorio-estadistico/monografia-de-bogota-2017\%5D
- http://www.sdp.gov.co/gestion-estudios-estrategicos/informacion-cartografia-y-estadistica/repositorio-estadistico/monografia-de-bogota-2011\%5D
SDM:
- http://www.sdp.gov.co/sites/default/files/determinantes_de_la_distribucion_espacial_de_informalidad_laboral_en_bogota.pdf

## Author Contributions

Conceptualization, J.P. and R.M.; methodology, J.P. and J.G.; software, J.P.; validation, R.M., J.G., and E.L.; formal analysis, J.P., J.G., and E.L.; investigation, J.P.; resources, J.P.; data curation, J.P. and E.L.; writing—original draft preparation, J.P.; writing—review and editing, J.P., R.M., J.G., and E.L.; visualization, J.P. and E.L.; supervision, R.M., J.G., and E.L.; project administration, J.G., and E.L. All authors have read and agreed to the published version of the manuscript.

## Footnotes

1 Unicode emojis (book) are generated with Latex.

2 Urban Sector is a cartographic division created by the National Administrative Department of Statistics (DANE for its acronym in Spanish).

3 The Bototá’s maps were created using shapes provided by Secretaría Distrital de Planeación de Bogotá for UPZ and National Administrative Department of Statistics (DANE) for Urban sectors.

4 The proposed domains are used for the convenience of the reader and could change depending on the data analysis made in the geographic area. It helps the reader to associate vulnerability factors related. These domains do not influence the process of assigning vulnerability to a spatial unit.

5 KDE uses the Gaussian kernel for its estimations and Scott’s Rule for the *bandwidth* selection [43].

6 The class identifier 1, …, *k* for the clusters of the vulnerability indexes with different *k* partitions (*k* = 3 left, *k* = 5 medium, *k* = 10 right) does not be the same between models (i.e., the class identifier variate from index to index).

7 The real COVID-19 cases are available in the publicly available dataset provided by Observatorio de Salud de Bogotá. The map shows the concentrations of the cases in 1000 meters on December 30, 2020.

## References

1. Madhav, N., Oppenheim, B., Gallivan, M., Mulembakani, P., Rubin, E., Wolfe, N. Pandemics: risks, impacts, and mitigation. In Disease Control Priorities: Improving Health and Reducing Poverty. 3rd edition; The International Bank for Reconstruction and Development/The World Bank, 2017.

2. PAHO. Leadership during a pandemic: What your municipality can do, 2009.

3. Moon, S., Sridhar, D., Pate, M.A., Jha, A.K., Clinton, C., Delaunay, S., Edwin, V., Fallah, M., Fidler, D.P., Garrett, L., others. Will Ebola change the game? Ten essential reforms before the next pandemic. The report of the Harvard-LSHTM Independent Panel on the Global Response to Ebola. The Lancet 2015, 386, 2204–2221.

4. Pathmanathan, I., O’Connor, K.A., Adams, M.L., Rao, C.Y., Kilmarx, P.H., Park, B.J., Mermin, J., Kargbo, B., Wurie, A.H., Clarke, K.R. Rapid assessment of Ebola infection prevention and control needs—six districts, Sierra Leone, October 2014. MMWR. Morbidity and mortality weekly report 2014, 63, 1172.

5. Lederberg, J., Hamburg, M.A., Smolinski, M.S., others. Microbial threats to health: emergence, detection, and response; National Academies Press, 2003.

6. Ethelberg, S., Lisby, M., Vestergaard, L., Enemark, H.L., Olsen, K., Stensvold, C., Nielsen, H., Porsbo, L.J., Plesner, A.M., Mølbak, K. A foodborne outbreak of Cryptosporidium hominis infection. Epidemiology & Infection 2009, 137, 348–356.

7. Ethelberg, S., Smith, B., Torpdahl, M., Lisby, M., Boel, J., Jensen, T., Nielsen, E.M., Mølbak, K. Outbreak of non-O157 Shiga toxin-producing Escherichia coli infection from consumption of beef sausage. Clinical infectious diseases 2009, 48, e78–e81.

8. Whittaker, P., Sopwith, W., Quigley, C., Gillespie, I., Willshaw, G., Lycett, C., Surman-Lee, S., Baxter, D., Adak, G., Syed, Q. A national outbreak of verotoxin-producing Escherichia coli O157 associated with consumption of lemon-and-coriander chicken wraps from a supermarket chain. Epidemiology & Infection 2009, 137, 375–382.

9. de Mattos Almeida, M.C., Caiaffa, W.T., Assunçao, R.M., Proietti, F.A. Spatial vulnerability to dengue in a Brazilian urban area during a 7-year surveillance. Journal of Urban Health 2007, 84, 334–345.

10. Moore, M., Gelfeld, B., Adeyemi Okunogbe, C.P. Identifying future disease hot spots: infectious disease vulnerability index. Rand health quarterly 2017, 6.

11. Salas, J., Yepes, V. Urban vulnerability assessment: Advances from the strategic planning outlook. Journal of Cleaner Production 2018, 179, 544–558.

12. UN Habitat. UN-habitat COVID-19 response plan, 2020.

13. WHO. Coronavirus disease 2019 (COVID-19) Situation Report - 40, 2020.

14. Mitlin, D. Dealing with COVID-19 in the towns and cities of the global South. IIED Blogs 2020, 27.

15. Hagenlocher, M., Kienberger, S., Lang, S., Blaschke, T. Implications of spatial scales and reporting units for the spatial modelling of vulnerability to vector-borne diseases. GI_Forum 2014, 2014, 197.

16. Kienberger, S., Hagenlocher, M. Spatial-explicit modeling of social vulnerability to malaria in East Africa. International journal of health geographics 2014, 13, 1–16.

17. Hagenlocher, M., Castro, M.C. Mapping malaria risk and vulnerability in the United Republic of Tanzania: a spatial explicit model. Population health metrics 2015, 13, 1–14.

18. Mishra, S.V., Gayen, A., Haque, S.M. COVID-19 and urban vulnerability in India. Habitat international 2020, 103, 102230.

19. Whitaker, R. Criticisms of the Analytic Hierarchy Process: Why they often make no sense. Mathematical and Computer Modelling 2007, 46, 948–961.

20. Mu, E., Pereyra-Rojas, M. Practical decision making: an introduction to the Analytic Hierarchy Process (AHP) using super decisions V2; Springer, 2016.

21. Chen, M.I., Lee, V.J., Barr, I., Lin, C., Goh, R., Lee, C., Singh, B., Tan, J., Lim, W.Y., Cook, A.R., others. Risk factors for pandemic (H1N1) 2009 virus seroconversion among hospital staff, Singapore. Emerging infectious diseases 2010, 16, 1554.

22. Jordan, R.E., Adab, P., Cheng, K. Covid-19: risk factors for severe disease and death, 2020.

23. Jung, S.m., Akhmetzhanov, A.R., Hayashi, K., Linton, N.M., Yang, Y., Yuan, B., Kobayashi, T., Kinoshita, R., Nishiura, H. Real-time estimation of the risk of death from novel coronavirus (COVID-19) infection: inference using exported cases. Journal of clinical medicine 2020, 9, 523.

24. Oppenheim, B., Gallivan, M., Madhav, N.K., Brown, N., Serhiyenko, V., Wolfe, N.D., Ayscue, P. Assessing global preparedness for the next pandemic: development and application of an Epidemic Preparedness Index. BMJ global health 2019, 4, e001157.

25. Morse, S.S. Factors in the emergence of infectious diseases. In Plagues and politics; Springer, 2001; pp. 8–26.

26. Summers, J.A., Wilson, N., Baker, M.G., Shanks, G.D. Mortality risk factors for pandemic influenza on New Zealand troop ship, 1918. Emerging infectious diseases 2010, 16, 1931.

27. Xu, B., Kraemer, M.U., Group, D.C. Open access epidemiological data from the COVID-19 outbreak. The Lancet. Infectious Diseases 2020.

28. Acharya, R., Porwal, A. A vulnerability index for the management of and response to the COVID-19 epidemic in India: an ecological study. The Lancet Global Health 2020, 8, e1142–e1151.

29. Grus, J. Data science from scratch: first principles with python; O’Reilly Media, 2019.

30. Angus, J.E. The probability integral transform and related results. SIAM review 1994, 36, 652–654.

31. Emerson, P. The original Borda count and partial voting. Social Choice and Welfare 2013, 40, 353–358.

32. SDP. Secretaría Distrital de Planeación: Monografías de las localidades Bogotá D.C. 2011, 2011.

33. SDP. Secretaría Distrital de Planeación: Monografías de las localidades Bogotá D.C. 2017, 2017.

34. SDM. Secretaría Distrital de Movilidad: Observatorio de Movilidad Bogotá D.C. 2017, 2018.

35. DANE. Departamento Administrativo Nacional de Estadística: COLOMBIA - Censo Nacional de Población y Vivienda - CNPV - 2018, 2018.

36. DANE. Departamento Administrativo Nacional de Estadística:: Pobreza multidimensional en Colombia, 2018.

37. DANE. Departamento Administrativo Nacional de Estadística: Índice de vulnerabilidad por manzana con el uso de variables demográficas y comorbilidades, 2020.

38. ILO. COVID-19 crisis and the informal economy: immediate responses and policy challenges 2020.

39. Goldstein, E., Lipsitch, M. Temporal rise in the proportion of younger adults and older adolescents among coronavirus disease (COVID-19) cases following the introduction of physical distancing measures, Germany, March to April 2020. Eurosurveillance 2020, 25, 2000596.

40. Alfonso R Ó.A. Densidades divergentes y segregación socio-espacial en la Bogotá de hoy: un análisis a partir de un índice de calidad de la densidad. VIII Seminario Internacional de Investigación en Urbanismo, Barcelona-Balneário Camboriú, Junio 2016. Departament d’Urbanisme i Ordenació del Territori. Universitat Politècnica, 2016.

41. SDP. Secretaría Distrital de Planeación: Determinantes de la distribución espacial de la informalidad laboral en Bogotá, 2018.

42. Gomez, J., Prieto, J., Leon, E., Rodriguez, A. INFEKTA: A General Agent-based Model for Transmission of Infectious Diseases: Studying the COVID-19 Propagation in Bogotá-Colombia. medRxiv 2020.

43. Scott, D.W. Multivariate density estimation: theory, practice, and visualization; John Wiley & Sons, 2015.

